# Survival and predictors of mortality after completion of TB treatment among people living with HIV

**DOI:** 10.1101/2022.05.18.22275233

**Authors:** Ivan Lumu, Joseph Musaazi, Aggrey Semeere, Ian Handel, Barbara Castelnuovo

## Abstract

**Background:** After completion of TB treatment patients may remain at-risk of complications and mortality. We determined the survival and predictors of all-cause mortality after completing TB treatment among ART experienced patients.

**Methods:** This was a retrospective cohort analysis of all ART experienced patients who completed TB treatment at a specialist HIV clinic in Uganda, between 2009 and 2014. The patients were followed for five years after TB treatment. We determined mortality rate, probability of death, and predictors of all-cause mortality after TB treatment using Poisson methods, Kaplan-Meier methods, and Cox proportional hazard models, respectively.

**Results:** A total 1,287 patients completed TB treatment between 2009 and 2014, of which 1,111 were included in the analysis. At TB treatment completion, the median age was 36 years (IQR: 31-42), 563 (50.7%) were males, and median CD4 count was 235 cells/mL (IQR: 139-366). The person time at risk was 4410.60 person-years. The all-cause mortality rate was 15.42 per 1000 person-years (95% CI: 12.14-19.59). The probability of death at five years was 6.9% (95%CI: 5.5%-8.8%). In the multivariable analysis, CD4 count<200 cells/mL was a predictor of all-cause mortality (aHR=1.81, 95%CI:1.06 - 3.11, *p*=0.03) alongside TB history (aHR=2.12, 95%CI: 1.16 - 3.85, *p*=0.01).

**Conclusion:** Survival post TB treatment in ART experienced PLHIV is reasonably good and most deaths occur within two years of TB treatment completion. Patients with low CD4 and those with history of treatment have an increased risk of mortality which underscores the need for TB prophylaxis, detailed assessment, and close monitoring after TB treatment.

**What is already known on this topic:** Tuberculosis is the leading cause of death in PLHIV and patients who complete treatment remain at risk of mortality. However, it is not clear what the mortality rate is, when it occurs, and what factors are associated with mortality in exclusively ART experienced patients.

**What this study adds:** Most deaths occur within two years after treatment completion decreasing drastically by year five. Patients with low CD4 count after TB treatment have an 81% increased risk of death and those with a history of TB have a 200% increased risk of mortality.

**How this study might affect research, practice or policy:** The study provides a detailed understanding of post-TB survival in ART experienced PLHIV and underscores the need for programs and clinics to re-define TB treatment success and consider the use of enhanced prophylaxis after TB treatment.

## INTRODUCTION

Tuberculosis(TB) is responsible for most AIDS-related deaths accounting for 214,000 deaths globally among People living with HIV (PLHIV) in 2020.^1^ People living with HIV are about five times more likely to develop TB as compared to HIV negative individuals, and the risk increases as the disease advances.^2, 3^ This risk, coupled with a high HIV prevalence in Sub Saharan Africa (SSA) is driving the TB epidemic further, despite the availability of effective TB treatment.^3^ Moreover, most TB deaths occur during the first three months of TB treatment initiation.^4^ Mortality during this period has been associated with low CD4 counts, not being on antiretroviral therapy (ART) ^5, 6^ and low body mass index (BMI)^4^ at the time of TB diagnosis, Some studies in HIV negative patients indicate that even after successful completion of TB treatment, up to 59% of patients get lung impairment,^7^ and consequently chronic lung disease.^8^ Moreover, some observational studies have reported reduced long term survival in TB survivors in the United State,^9^ and Vietnam. ^10^ The pooled standardised mortality after TB treatment in these studies is about three-fold higher than the general population.^11^ Death in these cohorts has been attributed to malignancies, cardiovascular complications, bacterial pneumonia, and TB reinfection.^10-12^ Similarly, reduced long term survival has been reported among PLHIV co-infected with TB at the time of entry in care in the UK, and Latin America.^13,14^ However, literature on long term survival after successful TB treatment in HIV and TB high burden settings is scarce. In the era of ART, such evidence on the long term prognosis and timing of these deaths would be important for HIV programs and clinics and would guide the designing of interventions to improve survival. Therefore, this study aimed to investigate the long-time survival of ART experienced patients and to determine the predictors of mortality after successful TB treatment.

## METHODS

### Study Design

We retrospective studied a cohort of all PLHIV who completed TB treatment between 1^st^ January 2009 and 31^st^ December 2014 at the Infectious Diseases Institute, Makerere University, Uganda.

#### Setting

The Infectious Diseases Institute clinic has registered over 33000 patients since its inception, and approximately 8000 patients are currently active in care. In 2009, the clinic adopted the WHO model of providing integrated care for HIV and TB in the same clinic to improve treatment outcomes.^15^ Patients’ care at the clinic follows the national TB and HIV treatment guidelines which are aligned with the WHO treatment guidelines. That is, an intensive phase of two months in new cases and a continuation phase of 4-6 months for drug-sensitive TB depending on the drugs used and the anatomic site of the disease. The quality control department regularly verifies the quality of patients’ records and updates the follow-up status of the patients. Patients’ follow-up status is maintained by active tracing and tracking of patients with missed appointments every two weeks.

#### Participant eligibility

This study included adults PLHIV aged ≥16 years who had completed TB treatment between 1^st^ January 2009 and 31^st^ December 2014. We excluded patients who were not taking lifelong ART during this period since lifelong ART is a known mortality modifier in PLHIV. ^16, 17^

#### Definition and procedures

Completion of treatment refers to any patient with clinical or microbiology evidence of tuberculosis that was started on anti-TB drugs and discharged as either ‘cured’ or ‘completed’ after taking the full course of the prescribed treatment. The date of TB treatment completion (discharge date) was used as the time of entry into the study. These patients were followed-up from the date of TB treatment completion to the date of the last visit in the event of death, transfer, or loss to follow up. Patients that were alive after five years were censored on the date of database closure.

#### Study endpoint

The primary end point was the time to all-cause mortality at 5-years after successful TB treatment. Secondary outcomes were; cumulative probability of death and mortality rate at one, three years and five years.

#### Statistical analysis

Descriptive analysis was done using frequencies and percentages for categorical variables or median and interquartile ranges (IQR) for continuous variables. Mortality rates were estimated using Poisson methods. Probabilities of deaths were estimated using Kaplan-Meier methods and compared using the Log-rank test. We examined the association between sex, age, calendar year of TB diagnosis, TB diagnosis method, previous TB history, TB type, WHO HIV clinical stage, and CD4 counts at TB treatment completion, Body Mass Index, ART duration, ART regimen type, and time to all-cause mortality after TB treatment using Cox proportional hazard regression model and report both unadjusted and adjusted Hazard ratios (uHR and aHR). All baseline characteristics (Table 1) were examined if they were associated with mortality. The factors which had a *p*-value<0.2 at unadjusted models were entered into the multivariable model, performed on complete cases, to adjust for confounding. However, each of the factors that had *p* value>0.2 at unadjusted analysis, were returned into the multivariable model one by one and then dropped if they had *p*-value>0.2. Statistical significance was set at a two-sided alpha of 0.05, and the *p*-value and 95% confidence interval is reported. We checked for; Cox proportional hazard assumption using Schoenfeld residuals, period effect by the inclusion of calendar year of TB diagnosis as a covariate in the Cox PH model; effect modification by fitting interactions between some covariates; multi-collinearity using variance inflation factor (VIF) method, and accounted for heterogeneity using robust standard errors in Cox PH model to account for variation across different calendar years. In the sensitivity analysis, the effect of loss to follow-up was examined using the worst-case scenario assumption that all patients lost to follow-up were dead, at time of loss to follow-up, as has been reported in some cohorts. ^18^ Also, sensitivity analysis was performed considering all patients lost to follow-up patients as alive and in care elsewhere at 5 years (best-case scenario). The effect of missing data on covariates was examined by refitting the final multivariable model using multiply imputed values from multiple imputations using chained equations (MICE) with 20 imputations, which were selected based on the proportion of missing values. All the statistical analysis was performed using STATA software (version 16.1).

**Table 1:** Participants’ characteristics at TB diagnosis or TB treatment completion.

| CHARACTERISTICS | NUMBER (%)<br>N=1,111 |
| --- | --- |
| **AT TB TREATMENT COMPLETION** |  |
| **SEX** |  |
| FEMALE | 548 (49.3) |
| MALE | 563 (50.7) |
| **AGE IN YEARS, MEDIAN (INTER-QUARTILE RANGE)** | 36 (31, 42) |
| **CATEGORIES** |  |
| 16_24 | 74 (6.7) |
| 25_34 | 396 (35.6) |
| 35_44 | 449 (40.4) |
| 45+ | 192 (17.3) |
| **CD4 COUNT (CELLS/ML) AT TB TREATMENT COMPLETION, MEDIAN (IQR)** | 235 (139, 366) |
| <200 | 393 (42.4) |
| >=200 | 533 (57.6) |
| **BODY MASS INDEX (KG/M<sup>2</sup>), MEDIAN (IQR)** | 22.0 (20.1, 24.2) |
| **CATEGORIES** |  |
| < 18 | 86 (8.2) |
| ≥ 18 | 964 (91.8) |
| **HIV WHO CLINICAL STAGE** |  |
| STAGE III | 463 (41.9) |
| STAGE IV | 642 (58.1) |
| **ART TYPE AT TB TREATMENT COMPLETION** |  |
| 1ST LINE | 1024 (92.2) |
| OTHER | 87 (7.8) |
| **ART DURATION IN YEARS, MEDIAN (IQR)** | 1.0 (1.0, 1.0) |
| **CATEGORIES** |  |
| 0 TO 1 | 856 (77.1) |
| 2 TO 4 | 137 (12.3) |
| ≥ 5 | 118 (10.6) |
| **AT TB DIAGNOSIS** |  |
| **CALENDAR YEAR OF TB DIAGNOSIS** |  |
| 2008_2010 | 376 (33.8) |
| 2011_2012 | 430 (38.7) |
| 2013_2014 | 305 (27.5) |
| **HEPATITIS B STATUS PRIOR/DURING TB TREATMENT** |  |
| NEGATIVE | 728 (94.3) |
| POSITIVE | 44 (5.7) |
| **TB TYPE** |  |
| PULMONARY TB | 649 (59.2) |
| EXTRA PULMONARY TB | 448 (40.8) |
| **TB DIAGNOSIS METHOD** |  |
| MICROBIOLOGICAL | 288 (25.9) |
| CLINICAL | 823 (74.1) |
| **TB HISTORY** |  |
| NO | 956 (86.0) |
| YES | 155 (14.0) |

Missing values: CD4 count (n=185), Body mass index (n=61), WHO stage (n=6), ART type (n=1).

## RESULTS

### Participants’ characteristics

A total of 1,287 PLHIV patients completed TB treatment from 2009 to 2014. Of these 166 were ART naive and 10 patients were on ART temporarily as prevention of mother to child transmission (PMTCT), and these were excluded from the analysis. A total of 1,111 patients were included in the analysis and Figure 1 shows the inclusion process and patients’ outcomes after follow-up. For the 1,111 patients, the median age was 36 years (IQR: 31-42), 51% were males, median CD4 counts was 235 cells/mL (IQR: 139 -366), and 393 (42%) had CD4 counts below 200 cells/ mL at TB treatment completion. Six hundred and forty-nine (59.2%) had been diagnosed with pulmonary TB, 155 (14.0%) had a history of previous TB treatment, and the majority (92.2%) were on first-line ART at TB treatment completion. The distributions of other patients’ characteristics are presented in Table 1.

**Figure 1:**
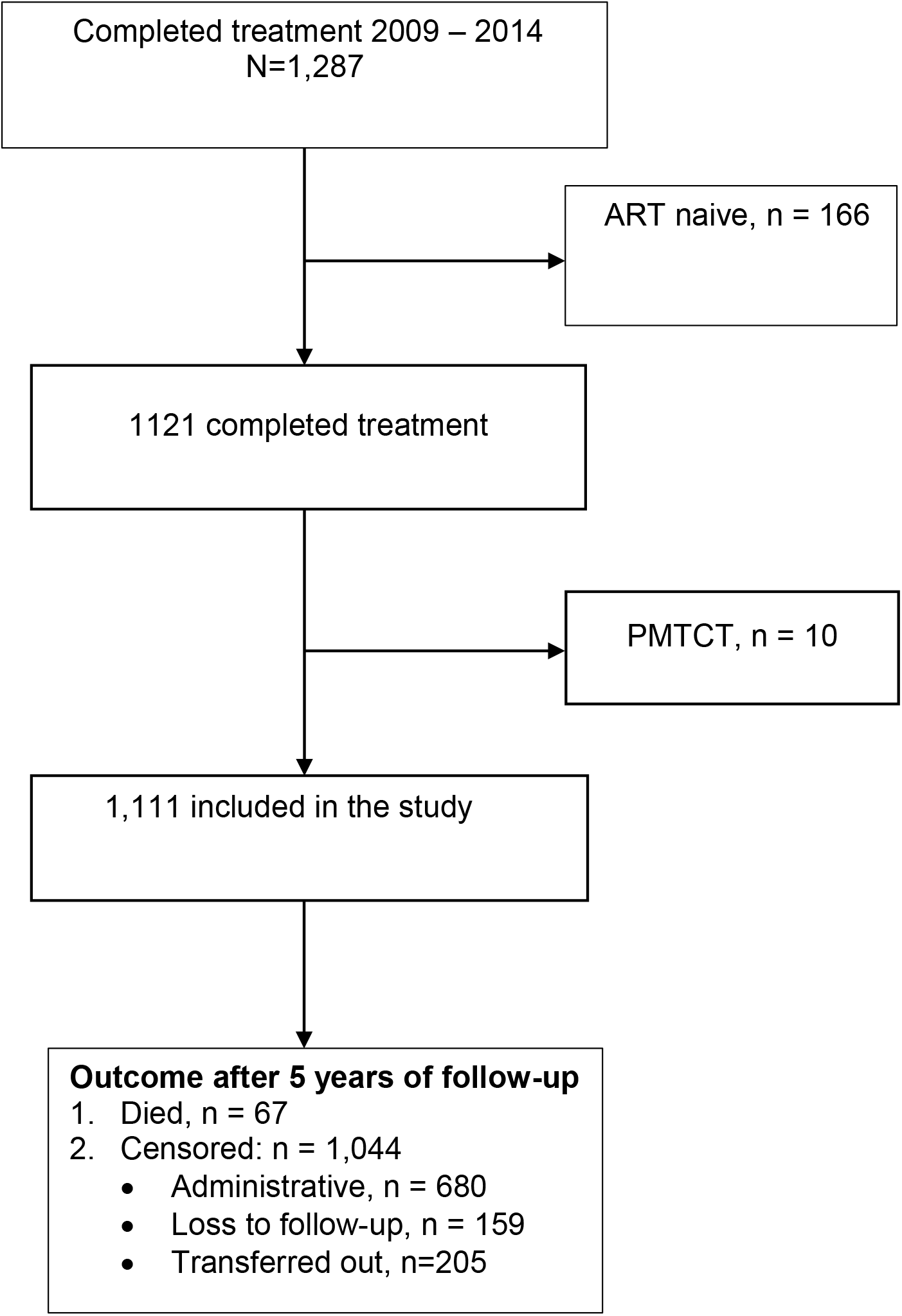
Patients’ inclusion process and outcomes during follow up.

### All-cause mortality post TB treatment

The average follow-up time from TB treatment completion was four years and the total person-time was 4,410.60 person-years (PY). Of the 1111 patients 159 (14.3%) were loss to follow-up, 205 (18.5%) were transferred-out, 67 (6%) died, and 680 (61.2%) were administratively censored. The overall all-cause mortality rate was 15.42 per 1000 person-years (95%CI: 12.14 - 19.59). The mortality at one, three, and five years was 25.89 per 1000 person-years (95%CI: 17.75 - 49.66), 10.65 per person-years (95%CI: 5.54 - 20.47), and 5.37 1000 person-years (95%CI: 2.02 -14.32), respectively (Figure 2). The rate of all-cause mortality varied by patient’s characteristic and was notably higher in patients with a body mass index <18Kgm^2^, <200cells/mL, 16-24 year-olds, Clinical Stage IV, and those with a history of TB treatment (Table S1). In the Kaplan-Meier analysis the cumulative probability of death at one year was 2.6% (95%CI: 1.8% - 3.7%), 5.8% (95%CI: 4.5% - 7.5%) at three years, and 6.9% (95%CI: 5.5%-8.8%) at five years (Figure 3A). By patient’s baseline characteristic, the probability of death was higher among patients with CD4 counts <200 cells/mL compared to those with CD4≥ 200 cells/mL (Log-rank test, *p*=0.01), and was higher among patients who had a previous history of TB compared to those with a first episode of TB (Log-rank test, *p*=0.02) (Figure 3B).

**Figure 2:**
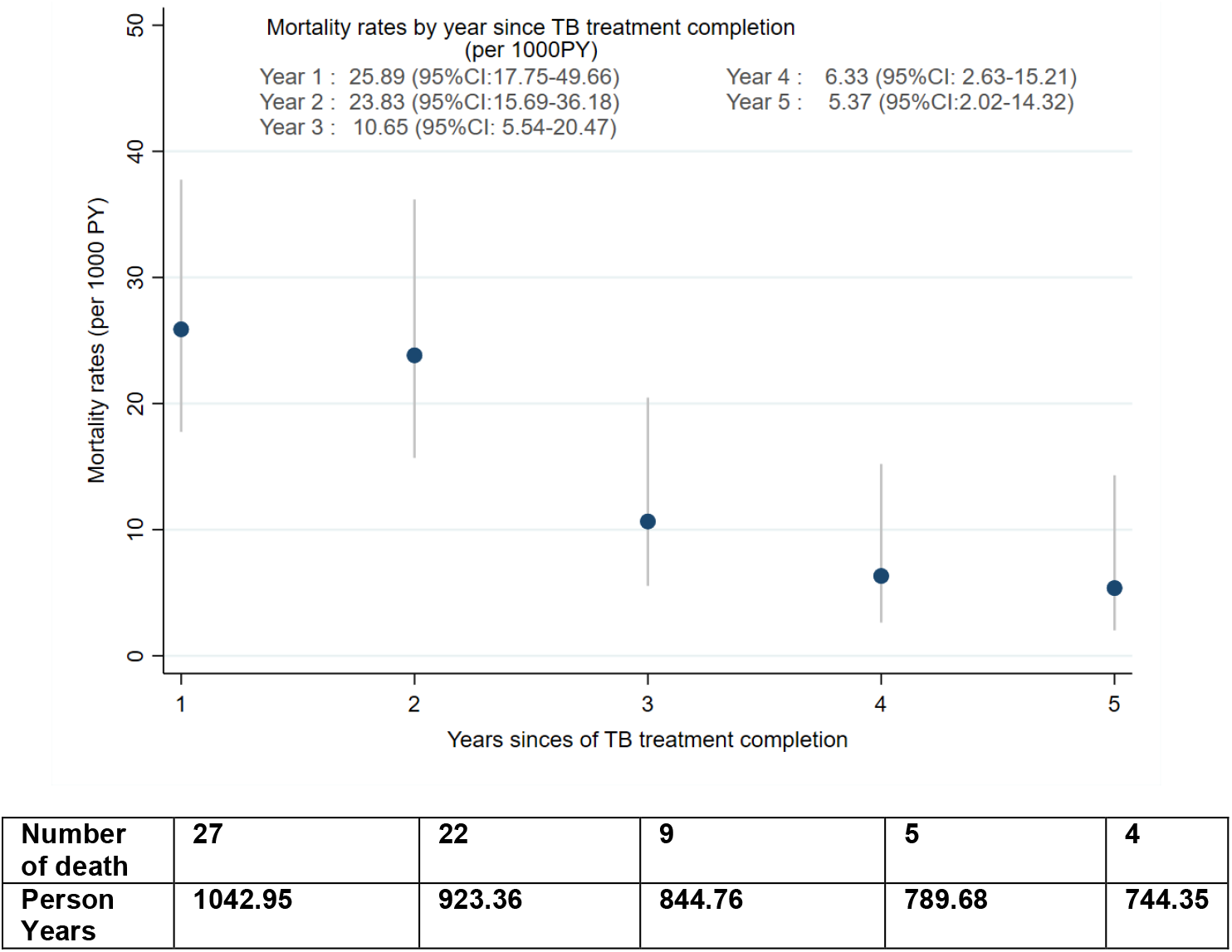
Mortality rates per 1000 Person-Years by year after TB treatment completion.

**Figure 3A:**
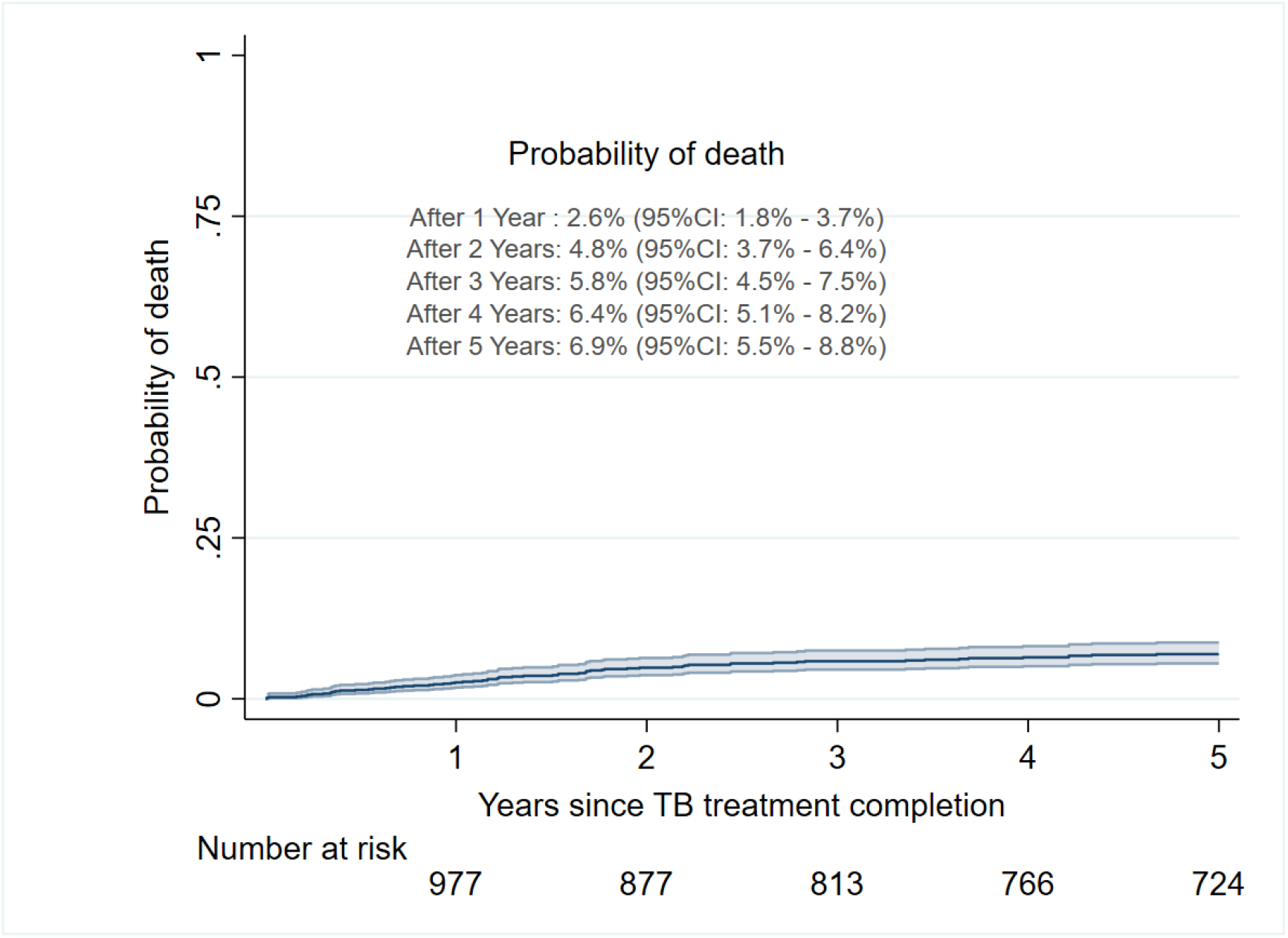
Kaplan-Meier for cumulative probability of death

**Figure 3B:**
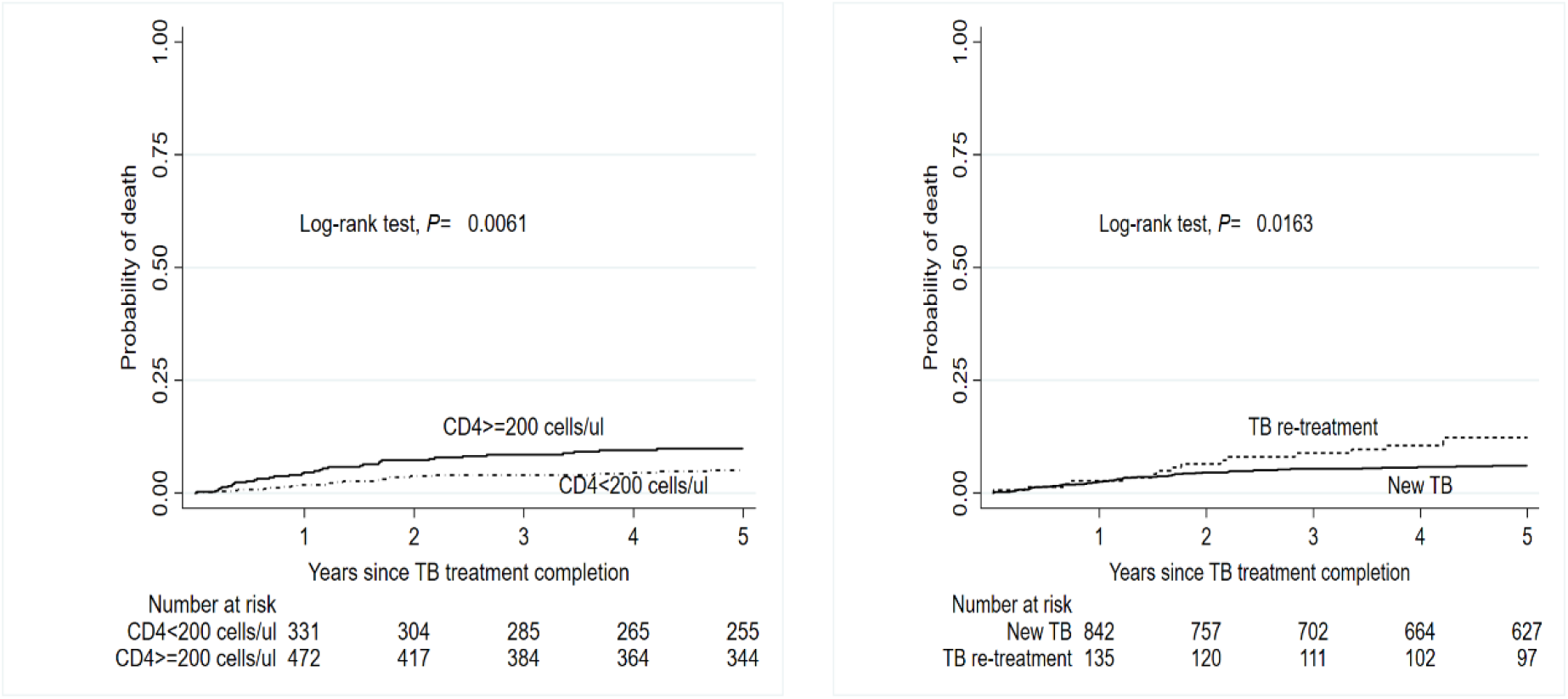
Kaplan-Meier for the cumulative probability of death by CD4 counts and TB patient type at TB treatment completion

In the sensitivity analysis, when all lost to follow-ups were considered as deaths, the mortality rate was 44.65 per 1000 person-years (95%CI 38.79-51.39) which is higher compared to the main analysis where losses to follow-up were censored (mortality rate 15.42 per 1000 PY (95%CI: 12.14 - 19.59). Similarly, the cumulative probability of death at five years was higher 19.2%, (95%CI: 17.0% - 21.7%) compared to the main analysis 6.9%(95%CI: 5.5% - 8.8%) when all lost to follow-up were considered dead (Figure S1). On the other hand, when patients lost to follow-up were considered alive but in care elsewhere the estimated mortality rate was similar to the main analysis at 14.14 per 1000 person-years with (95%CI 11.13-17.96).

### Factors associated with 5-years all-cause mortality post TB treatment

In the multivariable Cox proportional hazard regression model (Table 2), after adjusting for sex, calendar year of TB diagnosis, TB history, WHO HIV clinical stage and CD4 counts at TB treatment completion, CD4 count less than 200 cells/mL were significantly associated with death (aHR=1.81, 95%CI:1.06–3.11, *p* =0.03). Additionally, having previous TB history was significantly associated with death (aHR=2.12, 95%CI: 1.16-3.85, *p*=0.01). During the multivariable Cox PH regression, 17% (190/1,111) had missing data on at least one of the covariates in the adjusted model. After multiple imputations of missing data on covariates in the adjusted model, CD4 counts and TB history remained significantly associated with an increased risk of death (aHR=1.78, 95%CI: 1.06 -2.97, and 1.87, 95%CI: 1.06-3.30 respectively). Also, after multiple imputations, WHO HIV clinical stage IV at TB treatment completion became significantly associated with death (aHR=1.85, 95%CI: 1.08-3.17) (Supplementary, Table S2).

**Table 2:**
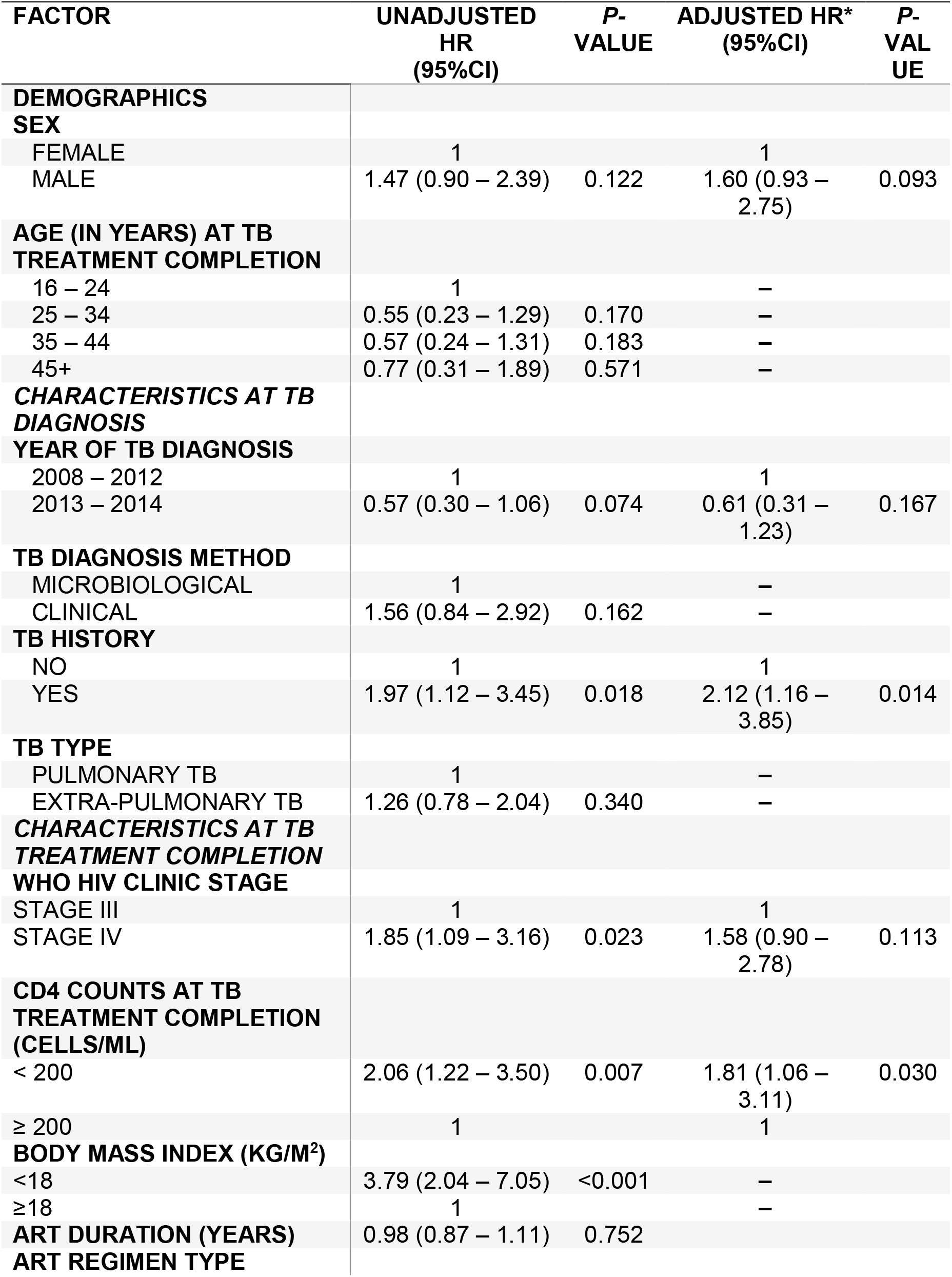
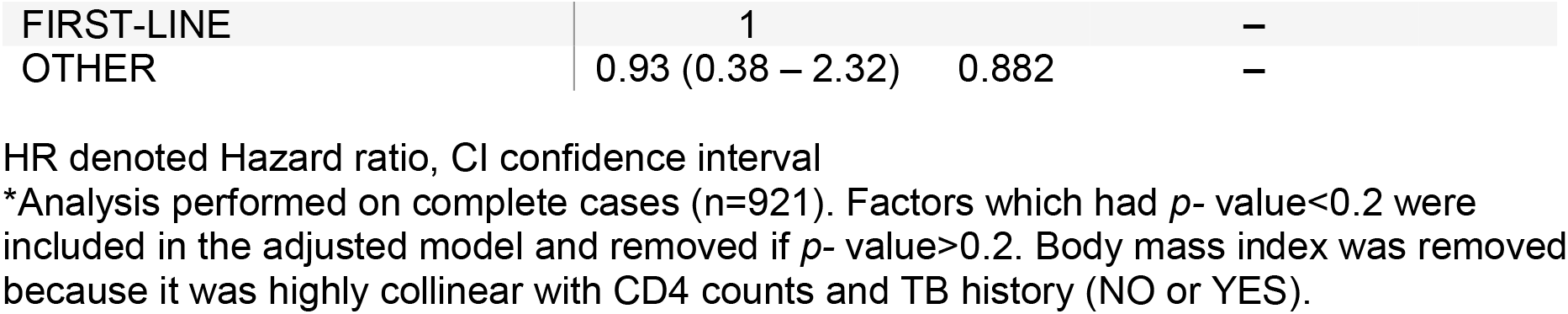
Cox proportional hazard model for mortality post TB treatment completion.

## DISCUSSION

To the best of our knowledge, this is the first study in sub-Saharan Africa to study the survival of ART experienced patients that complete TB treatment. In this cohort of ART experience patient mortality is highest in the first two years following treatment completion. It then decreased drastically and was lowest in the fifth year post TB. Similarly, the observed probability of death virtually doubled by year two and nearly plateaued after year three. The observation that most deaths post-TB treatment occur within two years of treatment completion could be attributed to; sequelae of severe TB disease, residual lung damage,^7^ recurrent bacterial infections post TB,^19^ and complications of poor immune recovery caused by HIV/TB coinfection.^20, 21^

Compared to other TB burden countries, the observed long term mortality rate (15.42 per 1000 person-years) was generally lower than has been previously reported in Ethiopia, and China.^22, 23^ Similarly, the adjusted 5-year mortality observation is lower than in HIV and TB co-infected cohorts in Latin America, and Myanmar. ^15, 24^ The better survival in our study may be attributed to IDI being a relatively well supported specialist referral clinic, early adoption of the WHO one-shop model of integrated services delivery,^15^ and recent advances in treatment and monitoring of PLHIV. On the other hand, the probability of death almost tripled at 5-years 19.2% (95%CI 17.0-21.8) when all lost to follow-up were considered dead in the sensitivity analysis, an indication that survival may be worse after TB treatment despite being on ART.

After adjusting for confounding in the multivariable Cox proportional hazard model, CD4 count<200 cells/mL was significantly associated with an 81% increased risk of death. This finding is consistent with the literature.^14^ The association with mortality could be that patients with a low CD4 count after TB treatment fail to achieve robust immune recovery despite being on ART, and thus remain susceptible to several opportunistic infections even with virological suppression.^21^ Furthermore, these patients may be more susceptible to complications from chronic inflammation due to TB and HIV replication. Such chronic inflammation leads to cardiovascular compilations which have been implicated as a cause of death post TB treatment.^10, 11^ Additionally, having a TB history was significantly associated with a 200% increased risk of death after TB treatment compared to those with no such history. This could probably be due the cumulative residual damage left by each TB episode in these patients,^25^ and hence a reduction in the survival.^10^ Additionally, TB recurrence in those with a history of TB could indicate ongoing immune dysfunction,^26, 27^ and hence an increased risk of mortality from AIDS and non-AIDS events in this sub-group. ^28^

The findings in this study are generalisable to HIV and TB programs worldwide and highlight the long term prognosis after completion of TB treatment in ART experienced patients. HIV programs and clinics should closely monitor patients in the first two years after treatment. During this period attention should be given to screening for AIDS events, cardiovascular and pulmonary complications. ^28, 29^ However, the implementation of such interventions means certain sub-groups will have to be prioritised. Such differentiated care should target those with malnutrition, immunological failure, and previous history of TB. These subgroups may require nutritional counselling and supplementation, use of enhanced prophylaxis,^30^ and further screening for opportunistic infections as part of the minimum package immediately after TB treatment. Additionally, the increased probability of death shortly after TB treatment coupled with the known risk of TB recurrence associated with low CD4 ^26^ underscore the need for clinics and programs to strengthen the coverage and uptake of intermittent preventive therapy(IPT) in TB survivors. Taken together, the results justify the need to redefined TB treatment success in persons living with HIV to include a time period and intervention package after TB treatment completion.

As it usually happens with retrospective studies, our study presents some limitations. There is a limited number of variables that could be explored with this clinical dataset. For instance, there was no data on socio-economic variables like education level, occupation status, alcohol use and homelessness which have been associated with long term mortality in some studies. ^12, 14^ Also, in resource-limited settings some lab tests are not regularly performed, like, haemoglobin and drug resistance testing (at the time of the study), yet anaemia and drug resistance may be associated with long term mortality. Lastly, the study did not establish the causes of death in these patients, and therefore it is impossible to quantify the contribution of post-TB disease. Despite the limitations, the study was sufficiently large with adequate observation and follow-up time of five years and included only patients on lifelong triple ART making the results generalisable to similar settings.

In conclusion, for ART experienced patients who complete TB treatment long-term survival is reasonably good, and mortality is highest within two years after treatment. Patients with low a CD4 cell count and those with a history of previous TB treatment have an increased risk of all-cause mortality and this underscores the need for details assessment, close monitoring, and the use of enhanced prophylaxis post TB treatment to improve survival. Further research is needed to understand the causes of death, and the contribution of post TB disease to morbidity and mortality.

## Data Availability

All data produced in the present study are available upon reasonable request to the authors

## Acknowledgement

We are grateful to the data management team that supported the abstraction of data.

## Contribution of the of the authors

LI designed the study and drafted the first manuscript.

LI and MJ analysed the data and reviewed the manuscript

SA: Reviewed the manuscript

HI Gave critical feedback on the design and data analysis and reviewed the manuscript.

CB supervised the design, data analysis, writing of the manuscript and reviewed the manuscript.

## Declaration of interest

L.I, M.J, S.A, H.I have no conflict of interest to declare.

CB is partly supported by the Fogarty International Centre, National Institute of Health (grant# 2D43TW009771-06 “HIV and co-infections in Uganda”).

## Data sharing agreement

The data collected including individual participant data will be made available to others as de-identified participant data. This data and statistical analysis plan will be accessed by emailing the corresponding author; however, some access criteria will be applied by the institution’s scientific research committee.

## Ethical considerations

At the infectious Diseases Institute, the use of de-identified secondary data is approved for analysis and publication by the institutional review board (IRB) of Makerere University College of Health Sciences (approval number: 120-2009). The IRB waived the need for informed consent to analyse this data.

## Role of funding

No funding was obtained to conduct this study. The corresponding author had access to all the data and had the final responsibility and decision to submit the results for publication.

## Notes

### Competing Interest Statement

The authors have declared no competing interest.

### Funding Statement

This study did not receive any funding

### Author Declarations

The institutional review board (IRB) of Makerere University College of Health Sciences gave ethical approval for this analysis (approval number: 120-2009), and the IRB waived the need for informed consent to analyse this data.

## REFERENCES

1. WHO. Global Tuberculosis Report 2021. Geneva: World Health Organisation, 2021

2. Gelaw YA, Williams G, Soares Magalhães RJ, Gilks CF, Assefa Y. HIV prevalence among tuberculosis patients in sub-Saharan Africa: a systematic review and meta-analysis. AIDS and Behavior. 2019 Jun;23(6):1561–75.

3. Bell LC, Noursadeghi M. Pathogenesis of HIV-1 and Mycobacterium tuberculosis co-infection. Nature Reviews Microbiology. 2018 Feb;16(2):80–90.

4. Musaazi J, Sekaggya-Wiltshire C, Kiragga AN, Kalule I, Reynolds SJ, Manabe YC, Castelnuovo B. Sustained positive impact on tuberculosis treatment outcomes of TB-HIV integrated care in Uganda. The International Journal of Tuberculosis and Lung Disease. 2019 Apr 1;23(4):514–21.

5. Ismail I, Bulgiba A. Predictors of death during tuberculosis treatment in TB/HIV co-infected patients in Malaysia. PLoS one. 2013 Aug 12;8(8):e73250.

6. Wondimu W, Dube L, Kabeta T. Factors Affecting Survival Rates Among Adult TB/HIV Co-Infected Patients in Mizan Tepi University Teaching Hospital, South West Ethiopia. HIV/AIDS (Auckland, NZ). 2020;12:157.

7. Pasipanodya JG, Miller TL, Vecino M, Munguia G, Garmon R, Bae S, Drewyer G, Weis SE. Pulmonary impairment after tuberculosis. Chest. 2007 Jun 1;131(6):1817–24.

8. Byrne AL, Marais BJ, Mitnick CD, Lecca L, Marks GB. Tuberculosis and chronic respiratory disease: a systematic review. International Journal of Infectious Diseases. 2015 Mar 1; 32:138–46.

9. Miller, T.L., Wilson, F.A., Pang, J.W., Beavers, S., Hoger, S., Sharnprapai, S., Pagaoa, M., Katz, D.J. and Weis, S.E., (2015) ‘Mortality hazard and survival after tuberculosis treatment’, American journal of public health, 105(5), pp. 930–937.

10. Fox GJ, Nguyen VN, Dinh NS, Nghiem LP, L. TN, Nguyen TA, Nguyen BH, Nguyen HD, Tran NB, Nguyen TL, L. TN. Post-treatment mortality among patients with tuberculosis: a prospective cohort study of 10 964 patients in Vietnam. Clinical Infectious Diseases. 2019 Apr 8;68(8):1359–66.

11. Romanowski K, Baumann B, Basham CA, Khan FA, Fox GJ, Johnston JC. Long-term all-cause mortality in people treated for tuberculosis: a systematic review and meta-analysis. The Lancet Infectious Diseases. 2019 Oct 1;19(10):1129–37.

12. Ranzani OT, Rodrigues LC, Bombarda S, Minto CM, Waldman EA, Carvalho CR. Long-term survival and cause-specific mortality of patients newly diagnosed with tuberculosis in São Paulo state, Brazil, 2010–15: a population-based, longitudinal study. The Lancet Infectious Diseases. 2020 Jan 1;20(1):123–32.

13. Zenner D, Abubakar I, Conti S, Gupta RK, Yin Z, Kall M, Kruijshaar M, Rice B, Thomas HL, Pozniak A, Lipman M. Impact of TB on the survival of people living with HIV infection in England, Wales and Northern Ireland. Thorax. 2015 Jun 1;70(6):566–73.

14. Koenig SP, Kim A, Shepherd BE, Cesar C, Veloso V, Cortes CP, Padgett D, Crabtree-Ramírez B, Gotuzzo E, McGowan CC, Sterling TR. Increased mortality after tuberculosis treatment completion in persons living with human immunodeficiency virus in Latin America. Clinical Infectious Diseases. 2020 Jun 24;71(1):215–7.

15. Hermans SM, Castelnuovo B, Katabira C, Mbidde P, Lange JM, Hoepelman AI, Coutinho A, Manabe YC. Integration of HIV and TB services results in improved TB treatment outcomes and earlier, prioritized ART initiation in a large urban HIV clinic in Uganda. Journal of acquired immune deficiency syndromes (1999). 2012 Jun 1;60(2):e29

16. Odone A, Amadasi S, White RG, Cohen T, Grant AD, Houben RM. The impact of antiretroviral therapy on mortality in HIV positive people during tuberculosis treatment: a systematic review and meta-analysis. PloS one. 2014 Nov 12;9(11): e112017.

17. Johnson LF, May MT, Dorrington RE, Cornell M, Boulle A, Egger M, Davies MA. Estimating the impact of antiretroviral treatment on adult mortality trends in South Africa: A mathematical modelling study. PLoS medicine. 2017 Dec 12;14(12): e1002468.

18. Brinkhof MW, Pujades-Rodriguez M, Egger M. Mortality of patients lost to follow-up in antiretroviral treatment programmes in resource-limited settings: systematic review and meta-analysis. PloS one. 2009 Jun 4;4(6): e5790.

19. Shuldiner J, Leventhal A, Chemtob D, Mor Z. Mortality after anti-tuberculosis treatment completion: results of long-term follow-up. The International Journal of Tuberculosis and Lung Disease. 2016 Jan 1;20(1):43–8.

20. Pawlowski A, Jansson M, Sköld M, Rottenberg ME, Källenius G. Tuberculosis and HIV co-infection. PLoS pathogens. 2012 Feb 16;8(2): e1002464.

21. Bisson GP, Zetola N, Collman RG. Persistent high mortality in advanced HIV/TB despite appropriate antiretroviral and antitubercular therapy: an emerging challenge. Current HIV/AIDS Reports. 2015 Mar;12(1):107–16.

22. Dangisso MH, Woldesemayat EM, Datiko DG, Lindtjørn B. Long-term outcome of smear-positive tuberculosis patients after initiation and completion of treatment: A ten-year retrospective cohort study. PloS one. 2018 Mar 12;13(3): e0193396.

23. Wang W, Zhao Q, Yuan Z, Zheng Y, Zhang Y, Lu L, Hou Y, Zhang Y, Xu B. Tuberculosis-associated mortality in Shanghai, China: a longitudinal study. Bulletin of the World Health Organization. 2015; 93:826–33.

24. Aung ZZ, Saw YM, Saw TN, Oo N, Aye HN, Aung S, Oo HN, Cho SM, Khaing M, Kariya T, Yamamoto E. Survival rate and mortality risk factors among TB–HIV co-infected patients at an HIV-specialist hospital in Myanmar: A 12-year retrospective follow-up study. International Journal of Infectious Diseases. 2019 Mar 1; 80:10-5.

25. Meghji J, Lesosky M, Joekes E, Banda P, Rylance J, Gordon S, Jacob J, Zonderland H, MacPherson P, Corbett EL, Mortimer K. Patient outcomes associated with post-tuberculosis lung damage in Malawi: a prospective cohort study. Thorax. 2020 Mar 1;75(3):269–78.

26. Lawn SD, Myer L, Edwards D, Bekker LG, Wood R. Short-term and long-term risk of tuberculosis associated with CD4 cell recovery during antiretroviral therapy in South Africa. AIDS (London, England). 2009 Aug 24;23(13):1717.

27. Kwan, C.K. and Ernst, J.D., 2011. HIV and tuberculosis: a deadly human syndemic. Clinical microbiology reviews, 24(2), pp. 351–376.

28. Pettit AC, Giganti MJ, Ingle SM, May MT, Shepherd BE, Gill MJ, Fätkenheuer G, Abgrall S, Saag MS, Del Amo J, Justice AC. Increased non-AIDS mortality among persons with AIDS-defining events after antiretroviral therapy initiation. Journal of the International AIDS Society. 2018 Jan;21(1): e25031.

29. Huaman MA, Kryscio RJ, Fichtenbaum CJ, Henson D, Salt E, Sterling TR, Garvy BA. Tuberculosis and risk of acute myocardial infarction: a propensity score-matched analysis. Epidemiology & Infection. 2017 May;145(7):1363–7.

30. Hakim J, Musiime V, Szubert AJ, Mallewa J, Siika A, Agutu C, Walker S, Pett SL, Bwakura-Dangarembizi M, Lugemwa A, Kaunda S. Enhanced prophylaxis plus antiretroviral therapy for advanced HIV infection in Africa. New England Journal of Medicine. 2017 Jul 20;377(3):233–45.

