## Supplementary material for "Survival and predictors of mortality after completion of TB treatment among people living with HIV": Table S1,Table S2,Figure S1

**TABLE OF CONTENTS**

**Table S1: Mortality rates by patients’ characteristics …………………………………………...1**

**Table S2: Cox proportional hazard model for mortality post TB treatment completion (on the imputation of missing values on covariates in the model) ………………………………………2**

**Figure 1: Kaplan-Meier for cumulative probability death assuming that all loss to follow-up were dead …………………………………………………………………………………………………3**

**Table S1: Mortality Rates by Patients’ Characteristics**

| **Characteristics** | **Person-time** | **Number of deaths** | **Mortality rate (per 1000 person-years)( 95% CI)** |
| --- | --- | --- | --- |
| ***At TB treatment completion*** |  |  |  |
| **Sex** |  |  |  |
| Female | 2162.37 | 27 | 12.49 (8.56- 18.21) |
| Male | 2182.74 | 40 | 18.33 (13.44- 24.98) |
| **Age in years, median (inter-quartile range)** |  |  |  |
| **Categories** |  |  |  |
| 16_24 | 281.93 | 7 | 24.83 ( 11.84- 52.08) |
| 25_34 | 1462.43 | 20 | 13.68 (8.82- 21.20) |
| 35_44 | 1797.15 | 25 | 13.91 (9.40- 20.59) |
| 45+ | 803.60 | 15 | 18.67 (11.25-30.96) |
| **CD4 count (cells/mL) at TB treatment completion, median (IQR)** |  |  |  |
| <200 | 1503.80 | 34 | 22.61(16.16- 31.64) |
| >=200 | 2074.87 | 23 | 11.09 (7.37- 16.68) |
| **Body mass index (Kg/m^2^), median (IQR)** |  |  |  |
| Categories |  |  |  |
| < 18 | 309.21 | 13 | 42.04(24.41-72.40) |
| ≥ 18 | 3898.53 | 43 | 11.03(8.18-14.87) |
| **HIV WHO clinical stage** |  |  |  |
| Stage III | 1863.15 | 19 | 10.20( 6.50- 15.99) |
| Stage IV | 2455.26 | 47 | 19.14(14.38- 25.48) |
| **ART type at TB treatment completion** |  |  |  |
| 1st line | 3992.11 | 62 | 15.53 (12.11-19.92) |
| Other | 353.00 | 5 | 14.16( 5.90- 34.03) |
| **ART duration in years, median (IQR)** |  |  |  |
| Categories |  |  |  |
| 0 to 1 | 3376.99 | 50 | 14.81(11.22- 19.54) |
| 2 to 4 | 504.67 | 50 | 19.81(10.66- 36.82) |
| ≥ 5 | 463.45 | 7 | 15.10( 7.20- 31.68) |
| ***At TB diagnosis/completion*** |  |  |  |
| **Calendar year of TB diagnosis** |  |  |  |
| 2008_2010 | 1432.96 | 22 | 15.35 (10.11- 23.32) |
| 2011_2012 | 1677.21 | 33 | 19.68(13.99 - 27.68) |
| 2013_2014 | 1234.94 | 12 | 9.72(5.52 -17.11) |
| **Hepatitis B status prior/during TB treatment** |  |  |  |
| Negative | 3342.25 | 18 | 5.39(3.39- 8.55) |
| Positive | 195.96 | 3 | 15.31 (4.94- 47.47) |
| **TB type** |  |  |  |
| Pulmonary TB | 2553.45 | 36 | 14.10 (10.17- 19.55) |
| Extra pulmonary TB | 1732.46 | 31 | 17.89 (12.58- 25.44) |
| **TB diagnosis method** |  |  |  |
| Microbiological | 1098.34 | 12 | 10.93 (6.20- 19.24) |
| Clinical | 3246.77 | 55 | 16.94(13.01- 22.06) |
| **TB history** |  |  |  |
| No | 3752.36 | 51 | 13.59(10.33- 17.88) |
| Yes | 592.75 | 16 | 26.99(16.54- 44.06) |

**Missing values: CD4 count (n=185), Body mass index (n=61), WHO stage (n=6), ART type (n=1),**

**Table S2: Cox proportional hazard model for mortality post TB treatment completion (on the imputation of missing values on covariates in the model)**

| **Factor** | **Unadjusted HR**  **(95%CI)** | **P-value** | **Adjusted HR***  **(95%CI)** | **P-value** |
| --- | --- | --- | --- | --- |
| **Demographics** |  |  |  |  |
| **Sex** |  |  |  |  |
| Female | 1 |  | 1 |  |
| Male | 1.47 (0.90 – 2.39) | 0.122 | 1.43 (0.87 – 2.33) | 0.158 |
| ***Characteristics at TB diagnosis*** |  |  |  |  |
| **Year of TB diagnosis** |  |  |  |  |
| 2008 – 2012 | 1 |  | 1 |  |
| 2013 – 2014 | 0.57 (0.30 – 1.06) | 0.074 | 0.65 (0.35 – 1.23) | 0.189 |
| **TB history** |  |  |  |  |
| No | 1 |  | 1 |  |
| Yes | 1.97 (1.12 – 3.45) | 0.018 | 1.87 (1.06 – 3.30) | 0.031 |
| ***Characteristics at TB treatment completion*** |  |  |  |  |
| **WHO HIV clinic stage** |  |  |  |  |
| Stage III | 1 |  | 1 |  |
| Stage IV | 1.85 (1.09 – 3.16) | 0.023 | 1.85 (1.08 – 3.17) | 0.024 |
| **CD4 counts at TB treatment completion (cells/mL)** |  |  |  |  |
| < 200 | 2.06 (1.22 – 3.50) | 0.007 | 1.78 (1.06 – 2.97) | 0.029 |
| ≥ 200 | 1 |  | 1 |  |

HR denoted Hazard ratio, CI confidence interval

*Analysis performed on multiply imputed values of missing values of covariates in the model (n=1,111). Thirty imputations were done with multiple imputation using chained equations.

Figure S1: Kaplan-Meier for cumulative probability **death** assuming that all loss to follow-up were dead

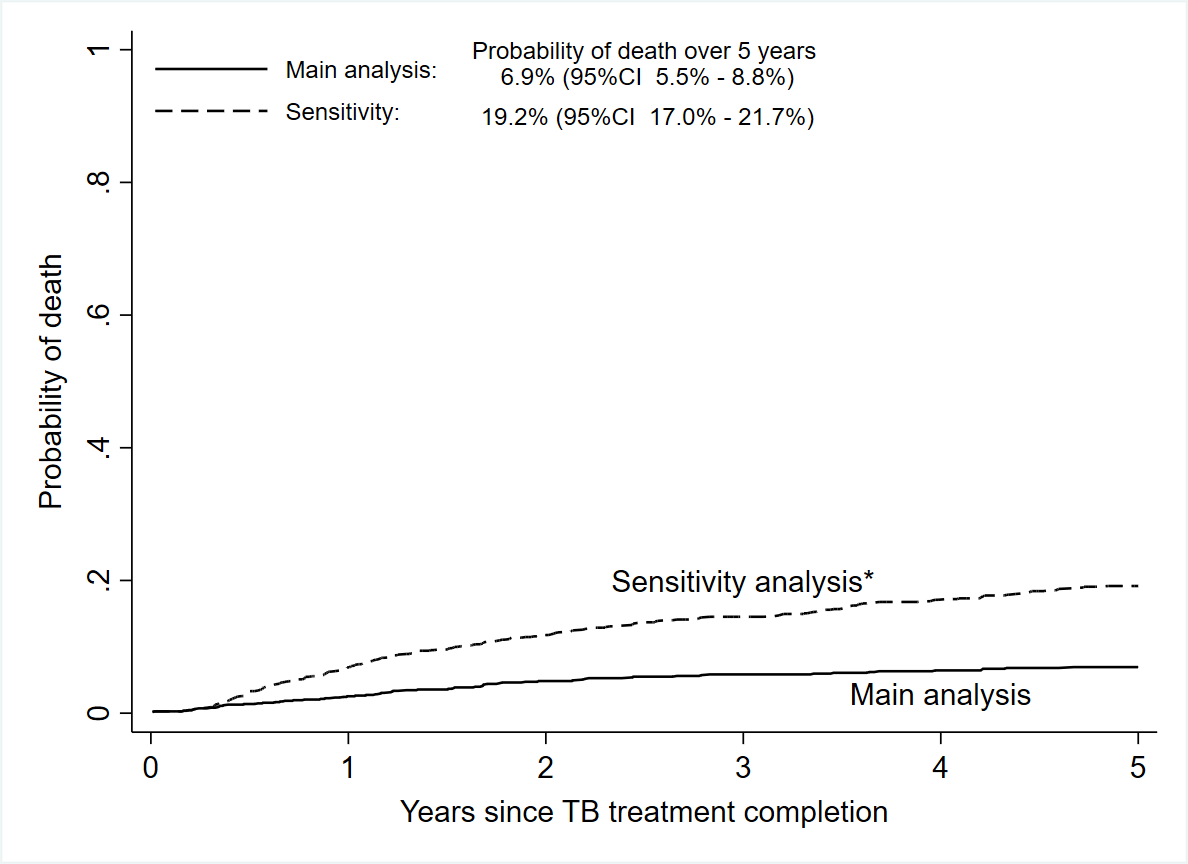
